# INTERMUSCULAR ADIPOSE TISSUE AND MUSCLE FUNCTION IN PATIENTS ON MAINTENANCE HEMODIALYSIS

**DOI:** 10.1101/2025.01.31.25321429

**Authors:** R. Gulsah Dilaver, Mert Demirci, Rachelle Crescenzi, Michael Pridmore, Lale A. Ertuglu, Andrew Guide, Robert Greevy, Baback Roshanravan, T. Alp Ikizler, Jorge L. Gamboa

**Author notes:** **Address correspondence to:** Jorge L. Gamboa, MD, PhD, University of Alabama at Birmingham, Division of Nephrology, Department of Medicine, Zeigler Medical Research Building, 627, 703 19th Street South, Birmingham, AL 35294.

## Abstract

**Background and Aims:** Sarcopenia, defined as a loss in muscle mass and strength, is common in patients with advanced chronic kidney disease (CKD), leading to poor outcomes. Intermuscular adipose tissue (IMAT) accumulation is associated with metabolic and functional abnormalities in chronic disease conditions. This study assesses IMAT in maintenance hemodialysis (MHD) patients and its association with metabolic markers and physical performance.

**Methods and Results:** We performed a cross-sectional study comparing MHD patients with controls. IMAT accumulation was measured by analyzing the fat-to-muscle ratio of the calf muscles through Magnetic Resonance Imaging (MRI) scans. Body composition and metabolic markers were assessed (hs-CRP, TNF-α, IL-6, and insulin resistance). Circulating cell-free mitochondrial DNA (ccf-mtDNA) was quantified using qRT-PCR. Muscle function was evaluated with handgrip strength. Inverse propensity weighted (IPW) method was used to test the difference between IMAT levels of the groups. Twenty-five MHD patients and 23 controls were analyzed. The MHD group had higher IMAT accumulation than controls (p < 0.01). IMAT was positively correlated with Body Mass Index (BMI) and fat mass index (FMI) in controls. MHD patients exhibited elevated TNF-α, IL-6, and hs-CRP levels (p < 0.01). Positive correlations were found between IMAT and IL-6 in MHD patients and between IMAT and TNF-α in controls. Handgrip strength was negatively correlated with IMAT in the entire cohort (p <0.01).

**Conclusion:** Our findings highlight the potential role of IMAT in muscle catabolism and functional decline in advanced CKD. Targeting IMAT could be a valuable strategy for improving health outcomes in this population.

## Introduction

Sarcopenia, defined as a loss in muscle mass and muscle strength, is common in patients with chronic kidney disease (CKD) and is strongly associated with poor physical function, morbidity, and mortality (1, 2). Multiple metabolic and nutritional abnormalities lead to sarcopenia in patients with CKD, including but not limited to the uremic state, systemic and local inflammation, insulin resistance, and metabolic acidosis (3-7).

Adipose tissue is a major endocrine organ in the body, producing and secreting hormones and pro-inflammatory cytokines that regulate metabolic systems (8). Subcutaneous adipose tissue (SAT), particularly in the thigh region, is considered protective against metabolic syndrome (9, 10). In contrast, ectopic or visceral adipose tissue (VAT) refers to fat accumulation in organs or tissues other than subcutaneous tissue, including muscle, and it is associated with adverse clinical outcomes (11). The presence and the extent of ectopic fat might be important in the systemic increase of circulating cytokines and contribute to inflammatory response in the tissues where they accumulate (12, 13). Higher Intermuscular adipose tissue (IMAT) accumulations are associated with several metabolic abnormalities, such as insulin resistance, systemic inflammation, and skeletal muscle mitochondrial dysfunction (14). IMAT accumulation is also associated with functional abnormalities in the skeletal muscle, such as low strength, quality, and activation, decreased gait speed, and decreased physical performance, which leads to increased frailty in affected individuals (15). Thus, the measurement of IMAT accumulation could provide insights into the development or worsening of sarcopenia through its paracrine communication (16, 17).

In this study, we aimed to determine the extent of IMAT accumulation in patients on maintenance hemodialysis (MHD) compared to individuals without kidney disease. We also examined the associations between IMAT accumulation and metabolic abnormalities commonly observed in advanced CKD, including systemic inflammation, insulin resistance, and physical performance. Our goal was to understand the impact of IMAT accumulation on overall health and to identify potential intervention targets to improve outcomes for individuals with advanced CKD.

## Methods

### Study Population

Participants were recruited from the Veteran Administration Nashville Campus (VANC) and the Vanderbilt University Medical (VUMC) Clinical Research Center between November 2018 and August 2023 as a part of an ongoing randomized clinical trial (NCT04067752). The primary inclusion criteria for the MHD group were treatment with a thrice-weekly outpatient hemodialysis program with single pool Kt/V greater than 1.2 for at least six months and well-functioning hemodialysis access. Individuals with eGFR higher than 60 ml/min/1.73 m^2^ were included as the control group. We recruited individuals aged 18 years or older. Exclusion criteria were active infectious or inflammatory disease (e.g., vascular access infections, active connective tissue disorders, active cancer, HIV, and liver disease), hospitalizations within the last month before the study, immunosuppressive agents (including high-potency topical steroids or prednisone at doses greater than 5 mg/day), and individuals with type 1 and type 2 diabetes mellitus who were using insulin or insulin-sensitizing medications. The Institutional Review Boards of VUMC and VANC approved the study protocol, and written informed consent was obtained from all study participants. The procedures were conducted under the principles outlined in the Declaration of Helsinki regarding the ethics of human research.

### Study protocol

Study procedures were performed at the Vanderbilt University Clinical Research Center (CRC) and Vanderbilt University Institute of Imaging Science. Participants initially underwent a standardized interview, a review of medical records, and a physical examination. Following an overnight 8-hour fasting period, they presented to CRC for blood draw and metabolic studies on a non-dialysis day. After metabolic studies, patients and controls underwent magnetic resonance imaging (MRI).

### Body Composition Measurement

The participants underwent a body composition analysis as part of a nutritional assessment. Body mass index (BMI) was utilized as the primary method for estimating body fat due to its widespread use (18). Fat mass index (FMI) was also determined using dual-energy X-ray absorptiometry (DXA) measurements by calculating the amount of fat mass in the body relative to height (kg/m^2^ ) to get a more accurate assessment of body composition (19).

### Magnetic Resonance Imaging

Subjects were placed in a 3.0-T MRI scanner (Philips Medical Healthcare, Andover, MA) in a supine position to image the leg at the widest part of the calf. The protocol involved the acquisition of five consecutive images from the leg, 10 mm thickness, and at intervals of 15 mm. Images from the leg were analyzed using software (MathWorks, Natick, MA) to calculate IMAT. The region of interest (ROI), which included the entire calf, was determined by manually tracing the fascia to exclude the bone and subcutaneous tissue. Signal intensities of the pixels in T1 weighted images were obtained to generate two distinct peaks that differentiate lean tissue from IMAT. IMAT was defined as fat beneath the deep fascia of the muscle and between muscle groups. The ratio of IMAT was calculated by dividing the IMAT cross-sectional area by the total muscle cross-sectional area.

### Metabolic Markers

After blood was drawn, samples were transported on ice and centrifuged at 3000 rpm for 15 minutes before being stored frozen at −80°C. Plasma fasting glucose concentrations were analyzed using the glucose oxidase method (Glucose analyzer 2; Beckman Coulter, Brea, CA). Biochemistry measurements were analyzed at the VUMC Pathology Laboratory. High-sensitivity C-reactive protein (hs-CRP) concentrations were measured using the high-sensitivity particle-enhanced turbidimetric UniCel DxI Immunoassay system (Beckman Coulter). Tumor necrosis factor alpha (TNF-α) and interleukin 6 (IL-6) were measured by enzyme-linked immunosorbent assay (R&D Systems, Minneapolis, MN). Insulin was measured by human radioimmunoassay (RIA) at Vanderbilt’s Hormonal Lab Core. Insulin resistance was calculated by the Homeostatic Model Assessment for Insulin Resistance (HOMA-IR).

### Circulating Cell-Free mtDNA

Circulating cell-free DNA (ccf-mtDNA) was measured as previously described (20) using plasma-stable DNA polymerase (Omni-KlenTaq-2 DNA Polymerase, DNA Polymerase Technology) and quantitative RT-PCR performed directly on patient plasma to quantify ccf-mtDNA. A PCR enhancer cocktail (PEC-2, DNA Polymerase Technology) was used to avoid inaccuracies associated with DNA purification methods. Primers specific for mitochondrially encoded NADH dehydrogenase 1 (ND1; forward 51- CCCTAAAACCCGCCACATCT-31and reverse 51- GAGCGATGGTGAGAGCTAAGGT-31) were used for ccf-mtDNA. Standard curves created with parallel quantitative RT-PCRs and oligonucleotide mimics of known concentrations were used to determine final DNA concentrations.

### Muscle Function Measurement

Muscle function was assessed using a handgrip test. Using a dynamometer, each participant’s handgrip strength was measured three times for both the right and left hands. The average handgrip strength for the right and left hands was calculated separately. The higher value of these averages among hands was then recorded as the participant’s handgrip strength in lbs.

### Statistical Analysis

This study had a cross-sectional design. The baseline characteristics of the participants were presented using the median and interquartile range (median, [IQR]) for age, BMI, FMI, and Systolic BP and the number and percentage for categorical variables such as gender, race, and diabetes history. Mann-Whitney U tests were used to compare non-parametric variables, and the Chi-Square test was used to compare categorical variables between the study groups. Spearman’s rank correlations were calculated to assess the relationship between each continuous variables of interest and the IMAT. Best-fitted line and confidence intervals were estimated using linear regression. Since our sample may not be adequately balanced across baseline demographics between the MHD and control groups, we conducted an inverse propensity weighted (IPW) method to test for a difference between the IMAT levels of the two groups. We balanced the cohort on age, gender, race, BMI, FMI, systolic blood pressure, and diabetes status. The threshold for statistical significance was set at p values ≤ 0.05. Analyses were performed using R Version 4.4.0.

## Results

### Demographics

A total of 650 patients were screened, and 35 patients consented to enroll in the MHD group. Of those, 31 patients were enrolled, and 25 of them had image compatibility, which refers to the quality and usability of the imaging data for IMAT calculation. A total of 350 participants were screened and 30 of them were consented to enroll in the control group. Twenty-five individuals were assessed for matching with MHD patients, and 23 had image compatibility and were included in the analysis **(Figure S1)**. Among the entire cohort, the median age was 53 (IQR, [47-66]) years, the body mass index (BMI) was 30.4 (IQR, [26.8 – 34.5]) kg/m^2^ , 83% were male, and 56% were African American. Three had a history of type 2 Diabetes Mellitus (DM), and all were off antidiabetic medications. The baseline characteristics were similar between MHD and control groups in terms of age, gender, and BMI, although African American participants were significantly higher in number in the MHD group **(Table 1)**.

**Table 1.**
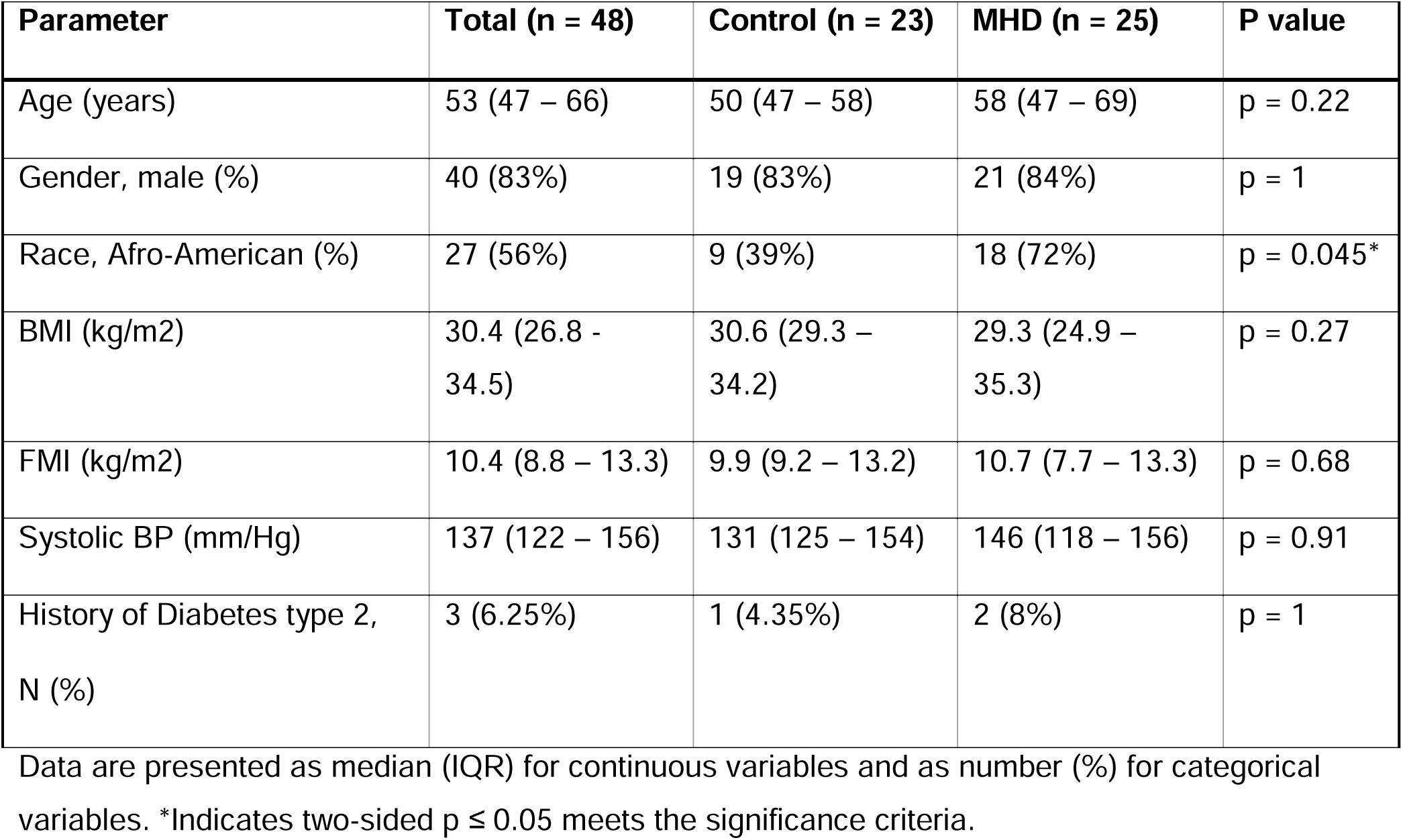
Demographic and Clinical Characteristics of Study Participants.

The baseline laboratory values are depicted in **Table S1.** As expected, serum hematocrit, total cholesterol, and HDL cholesterol levels were significantly lower in the MHD group (p < 0.01). Pre-albumin concentrations were higher in patients on MHD compared to controls with pre-albumin of 33.5 (IQR, [23.8 – 38.4]) mg/dl vs 24.3 (IQR, [22.1 – 26]), (p < 0.01), respectively. There were no significant differences between serum albumin and triglyceride concentrations between the study groups (p = 0.1, p = 0.8, respectively).

### IMAT Accumulation in Patients on MHD and Controls

IMAT level, expressed as the ratio of IMAT area to muscle area, was 0.0686 (IQR, [0.0601 – 0.1149]) in the entire cohort. Median IMAT was 0.113 (IQR, [0.065 – 0.146]) in patients treated with MHD versus 0.064 (IQR, [0.047 – 0.074]) in controls (p < 0.01). **(Figure 1A, 1B)**. After balancing across demographics, a weighted t-test on the weighted cohort from the IPW analysis showed IMAT levels 0.044 (95% CI: 0.021, 0.068) were higher in the MHD group compared to controls.

**Figure 1.**
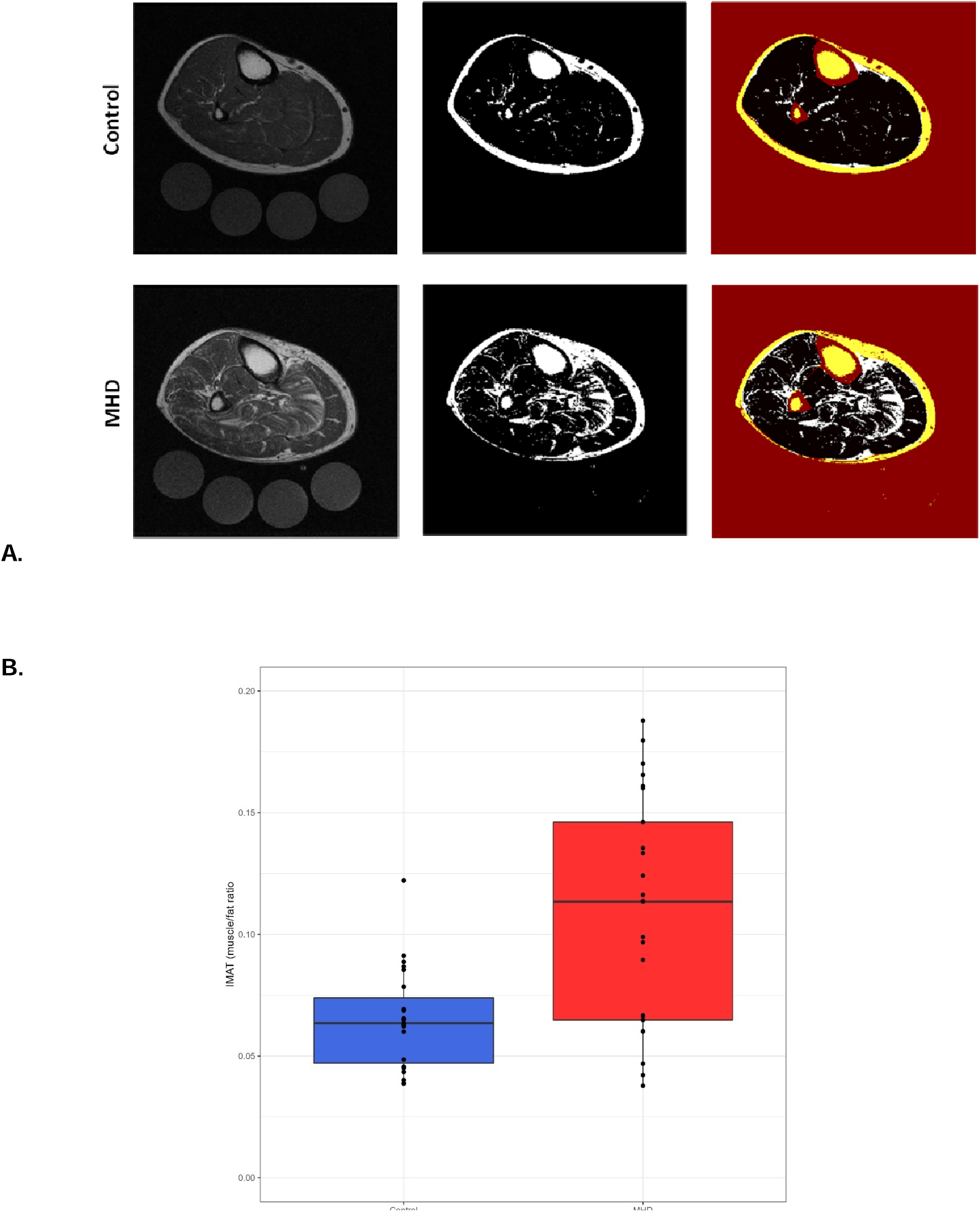
IMAT Accumulation in Participants. **(A)** Representative IMAT accumulation in the leg muscle cross-sectional area. A representative image of a control (top row) and a MHD (bottom row) subject. Images include MRI Dixon fat-weighted image, threshold determination for IMAT accumulation, and excluding the subcutaneous fat, skin, bone, and vascular part for calculation of IMAT accumulation ratio (red and yellow parts). **(B)** Box plot for IMAT content comparison between control and MHD groups (p = 0.001).

### Body Composition and IMAT Accumulation

There was no meaningful or statistically significant correlation between IMAT accumulation and BMI for the entire cohort (r = 0.28, p = 0.06, **Figure 2A)**. When the correlations were analyzed within each group, there was a statistically significant positive relationship between IMAT and BMI in the control group only (r = 0.54, p < 0.01, **Figure 2B, 2C)**. There was no statistically significant correlation between IMAT accumulation and FMI for the overall cohort (r = 0.26, p = 0.07). When analyzed separately by groups, a significant correlation was found between the IMAT accumulation and FMI in the control group (p < 0.05, r = 0.47) but not for the MHD group, **(Figure 2D, 2E, 2F)**.

**Figure 2.**
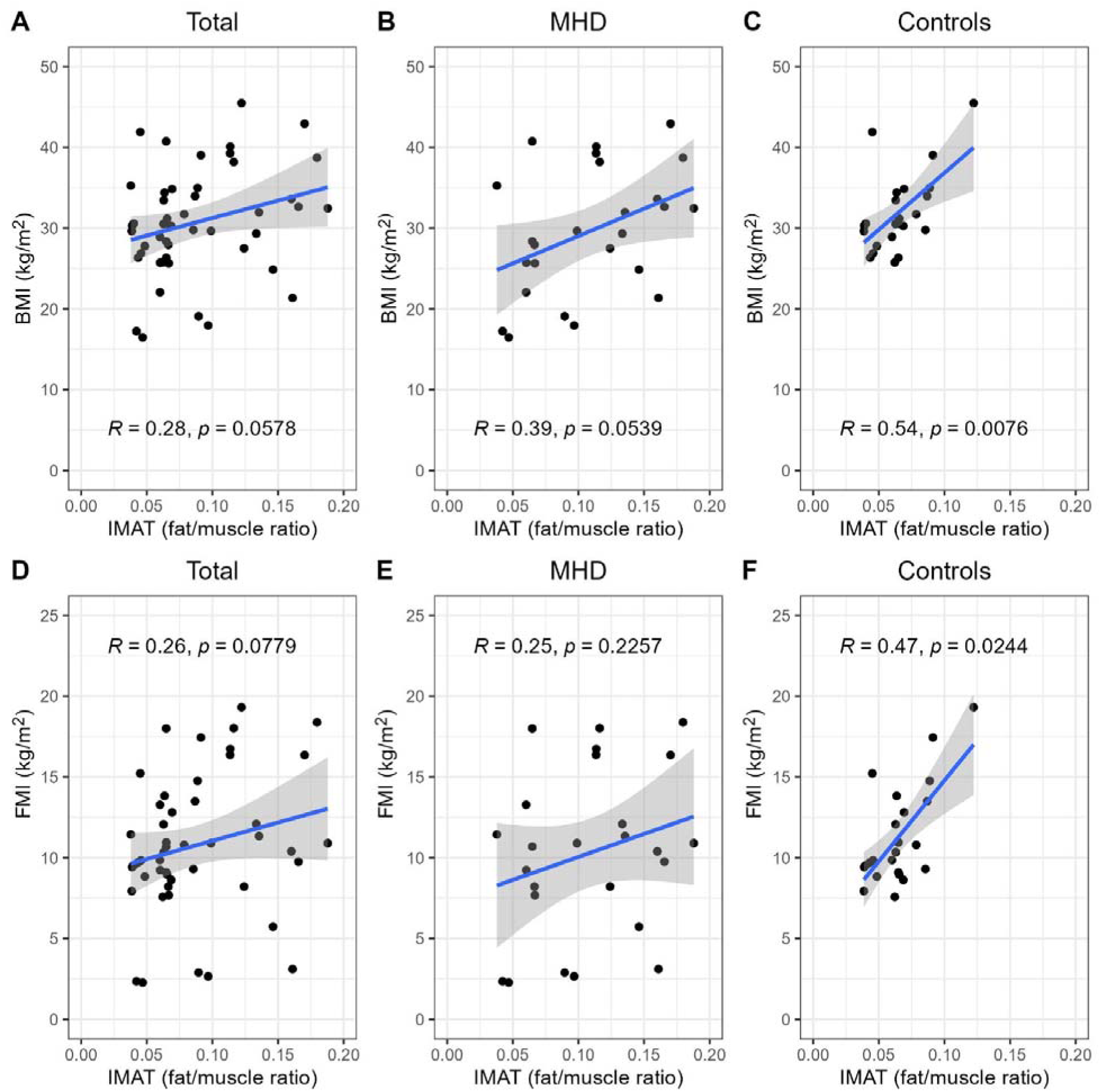
Correlation graphs of IMAT accumulation and body compositions parameters. The Spearman correlation analysis shows the relationship between IMAT accumulation and BMI and FMI for the entire cohort as well as separately for the MHD and control groups.

### Inflammatory Markers and IMAT Accumulation

**Table2** displays the comparisons of various inflammatory markers among groups. Significantly higher levels in patients undergoing MHD were observed for TNF-α, IL-6, and hs-CRP (p < 0.001), (**Figure S2A, S2B, S2C)**. Positive correlations were found for the entire cohort between TNF-α and IMAT (r = 0.41, p = 0.004), IL-6 and IMAT (r = 0.48, p < 0.001), and hs-CRP and IMAT (r = 0.48, p < 0.001), (**Figure 3A, 3D, 3G)**. When analyzed separately, there was a significant correlation between IMAT and IL-6 in the MHD group (r= 0.46, p < 0.05) but not in the control group **(Figure 3E, 3F**). There was no significant correlation between IMAT and hs-CRP in either group (**Figure 3H, 3I).** Additionally, there was a significant correlation between IMAT and TNF-α in the control group (r = 0.45, p < 0.05), but not in the MHD group **(Figure 3B, 3C)**.

**Figure 3.**
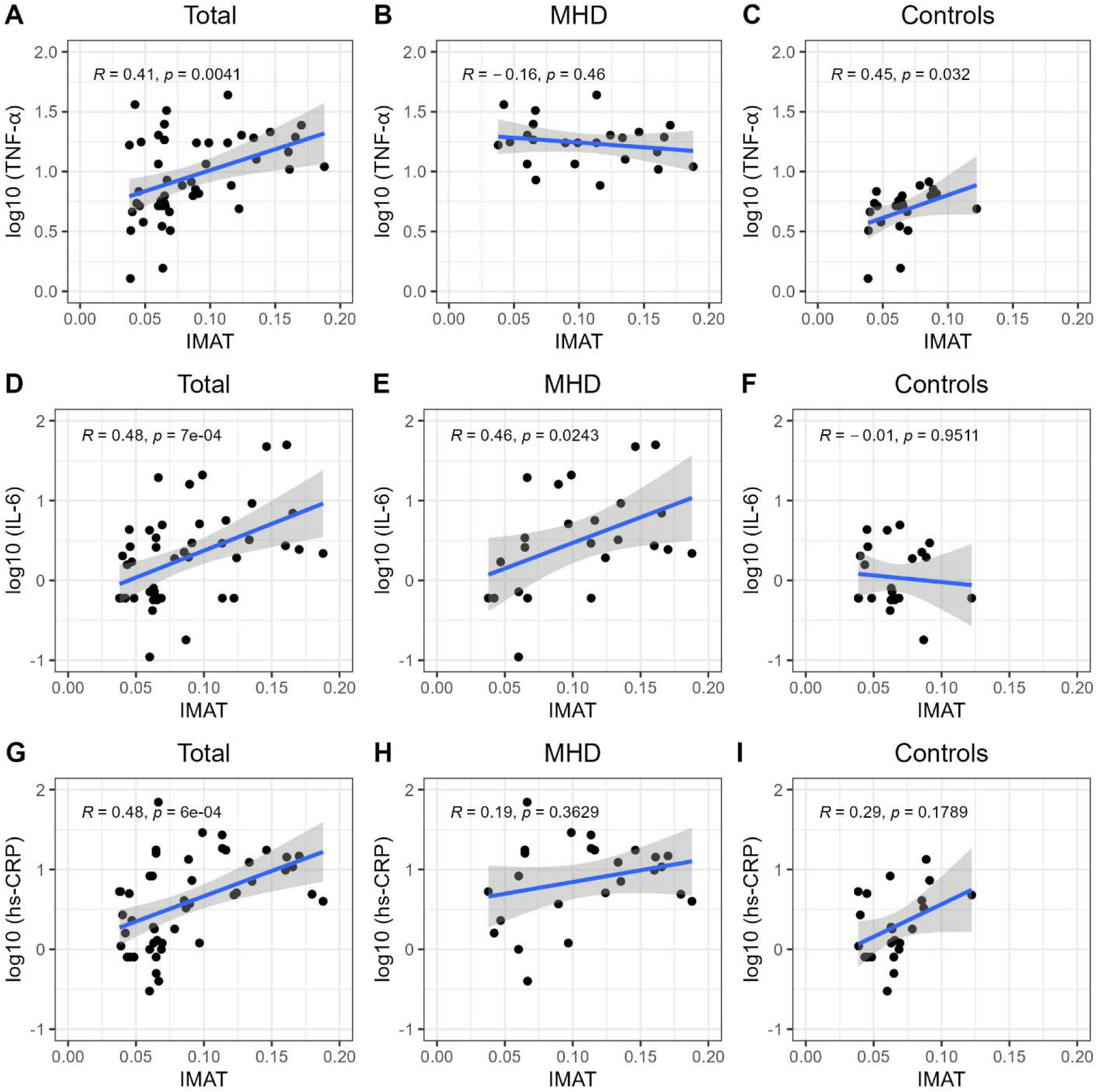
Correlation graphs of IMAT and inflammatory marker concentrations with their logarithmic transformations. The Spearman correlation analysis shows the relationship between IMAT accumulation and TNF-α, IL-6, and hs-CRP for the entire cohort as well as separately for the MHD and control groups.

### ccf-mtDNA and IMAT Accumulation

ccf-mtDNA levels were 9.47 (IQR, [6.88 – 20.15]) pM for the control group and 8.66 (IQR, [5.33 – 48.48]) pM for MHD participants and were not significantly different. There were no meaningful or significant correlations between ccf-mtDNA and IMAT results for the entire cohort (p = 0.19,) or within groups (p = 0.07 and p = 0.9, for the control and MHD groups, respectively). In addition, ccf-mtDNA did not correlate with any of the inflammatory markers.

### Insulin Resistance and IMAT Accumulation

Higher HOMA-IR levels were observed in patients undergoing MHD compared to controls [4.51 (IQR, [2.49 – 6.00]); versus 3.11, (IQR, [2.38 – 4.31]), respectively], although the difference was not statistically significant (p = 0.2). There were no statistically significant correlations between IMAT accumulation and HOMA-IR for the entire cohort (p = 0.18) or in subgroup analysis.

### Handgrip Strength and IMAT Accumulation

Maximum handgrip strength was significantly lower in the MHD group versus the control group [63 (IQR, [55 – 77]) lbs. versus 93 (IQR, [73.5 – 104.5]) lbs. respectively, p < 0.01], **(Table 2), (Figure 4A)**. There was a negative correlation between handgrip strength and IMAT for the entire cohort (r = - 0.38, p < 0.01), **(Figure 4B).** There were no significant correlations when MHD and control groups were analyzed separately (**Figure 4C, 4D)**.

**Figure 4.**
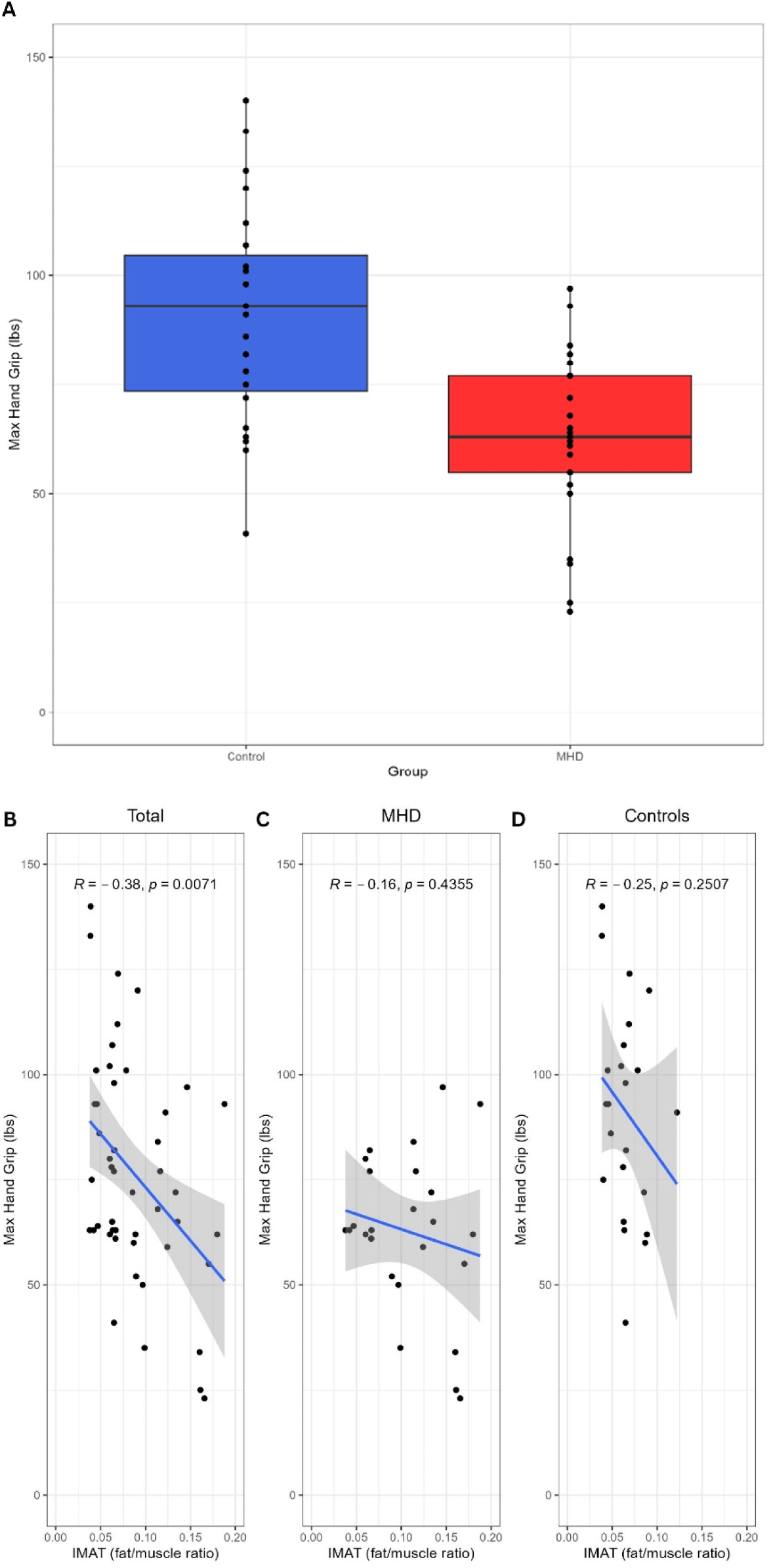
Comparison of IMAT accumulation and hand grip strength between groups. **(A)** Box plot for handgrip test comparison between control and MHD groups (p<0.05). The Spearman correlation analysis shows the relationship between IMAT accumulation and handgrip strength **(B), (C), (D).**

**Table 2.** Inflammatory Markers, and Other Outcomes.

| Parameter | Total (n = 49) | Control (n = 23) | MHD (n = 25) | P - value |
| --- | --- | --- | --- | --- |
| IL-6 (pg/mL) | 1.95 (0.60 – 3.84) | 0.72 (0.60 – 2.14) | 2.82 (1.46 – 7.54) | P = 0.004* |
| TNF-α (pg/mL) | 8.22 (5.17 – 17.53) | 5.17 (4.20 – 6.28) | 17.52 (12.37 – 20.48) | P < 0.001* |
| hs-CRP (mg/L) | 4.5 (1.2 – 11.2) | 1.8 (0.9 – 4.4) | 9.8 (4.0 – 17.5) | P < 0.001* |
| HOMA-IR | 3.72 (2.38 – 5.39) | 3.11 (2.38 – 4.31) | 4.51 (2.49 – 6.00) | P = 0.21 |
| mtDNA | 9.41 (6.88 – 20.15) | 9.47 (8.33 – 12.14) | 8.66 (5.33 – 48.48) | P = 1 |
| Maximum Handgrip Strength (lbs) | 73.5 (62.0 – 93.0) | 93.0 (73.5 – 104.5) | 63.0 (55.0 – 77.0) | P < 0.001* |

Data are presented as median (IQR). \*Indicates two-sided $p \leq 0.05$ meets the significance criteria.

## Discussion

In this cross-sectional study, we found that patients on MHD had significantly higher IMAT levels compared to individuals without kidney disease. The propensity weighting also controls for potential confounding in the data and provides strong evidence of a clear difference in IMAT levels between MHD and control patients. Significant correlations were identified between IMAT, body composition, and inflammatory biomarkers, such as TNF-α and IL-6. Additionally, there was a notable negative correlation between IMAT and handgrip strength, indicating impaired muscle function with higher IMAT levels.

As an ectopic fat depot, IMAT is a sign of chronic fatty accumulation, which worsens with many chronic diseases (13). While implications of increased IMAT are not fully elucidated, it is linked to muscle weakness in the setting of chronic diseases, which often accompanies sarcopenia (21). Increased levels of IMAT have also been linked to insulin resistance and inflammation, which can further contribute to muscle dysfunction, decreased mobility, and increased frailty (22-25). Our data are consistent with those previous reports with significantly increased IMAT in patients on MHD compared to matched controls. Given the high prevalence of sarcopenia and poor physical performance in patients with moderate to advanced CKD, understanding the extent and the role of IMAT accumulation in MHD patients is important for identifying potential therapeutic targets to improve patient outcomes.

In our study, a higher BMI and FMI were significantly associated with increased IMAT in the control group. This relationship in patients without CKD could be attributed to the general increase in body fat mass and a consequent increase in IMAT (17). Our results suggest that the mechanism of IMAT accumulation in patients on MHD could be different than those without CKD and not solely driven by increased body adiposity. In chronic conditions such as sarcopenia, greater IMAT is associated with greater inflammation, accelerated aging, uremia, reduced physical activity, and insulin resistance independent of overall adiposity (26).

The significant correlations between IMAT and inflammatory markers hs-CRP, IL-6, and TNF-α suggest that ectopic fat accumulation and inflammation share common mechanistic pathways. The pro-inflammatory nature of IMAT may play a key role in this relationship, as ectopic fat tissue contributes to elevated peripheral cytokine levels and exacerbates muscle catabolism and dysfunction through a paracrine effect (14). In this study, the discrepancy in several within-group correlations between MHD and controls is may be related to the complexity of the inflammatory responses and the interplay of multiple factors, particularly in patients with CKD. The wider range of inflammatory biomarker values in MHD patients may also suggest a greater heterogeneity of inflammatory phenotypes in the MHD. Further studies should evaluate the contribution of IMAT to the increased systemic inflammation in CKD.

It has been reported that high plasma ccf-mtDNA levels are associated with inflammation in various disease states (27, 28). The release of mtDNA into the plasma could be the result of mitochondrial dysfunction. We previously reported that mitochondrial dysfunction is present in patients with moderate to advanced CKD even before the initiation of maintenance dialysis. We have also found that muscle mitochondrial dysfunction is associated with IMAT accumulation in CKD (29). In our current study, we did not find any difference in ccf-mtDNA levels between study groups, and there was no association between ccf-mtDNA and IMAT. Also, there was no correlation between ccf-mtDNA and inflammatory markers. Further prospective studies that improve mitochondrial function or decrease IMAT are required to evaluate the relationship between these two variables in chronic kidney disease.

Multiple studies show increased ectopic fat accumulation linked to higher insulin resistance levels (30, 31). Ectopic fat accumulation increases the release of inflammatory cytokines and the entry of free fatty acids into the circulation, thereby reducing insulin’s ability to regulate glucose uptake. IMAT accumulation in specific muscle groups has been associated HOMA-IR in previous reports (32, 33). Although we observed a positive trend between IMAT and HOMA-IR, this was not statistically significant, which could be due to the relatively small sample size in our patient subgroups.

Handgrip strength is a simple measurement of maximum voluntary muscle strength and is used as a reliable tool for diagnosing sarcopenia and assessing overall muscle strength (34). In our study, we found a significant negative correlation between IMAT and handgrip strength, indicating that higher IMAT is associated with lower muscle strength. Previous studies have shown the association between IMAT and physical performance in the presence of IMAT around muscle fibers, which could interfere with muscle contraction and force, leading to lower muscle strength (35). Additionally, IMAT may trigger local inflammation and metabolic abnormalities in the muscle tissue, worsening muscle breakdown and hindering muscle repair. Alternatively, it is also possible that weakened muscles may lead to increased fat accumulation within the muscle. This bidirectional relationship highlights the need for further research to understand the underlying mechanisms.

Our study has several strengths. We assessed IMAT accumulation thoroughly and included multiple gold standard markers for assessment of body composition, muscle function, and inflammatory markers in the analyses. The study focuses on a high-risk population for sarcopenia and associated poor outcomes, providing insights that can help target future interventions for these patients. There are some limitations to consider. The small sample size at a single institution limits the power to detect specific associations and the generalizability of the findings. More importantly, the cross-sectional design precludes establishing causality between IMAT accumulation and the observed abnormalities. The lack of longitudinal data restricts understanding of progressive IMAT accumulation and its long-term effects. Other comorbidities in MHD patients could also influence the results, and not all confounding factors may have been accounted for. Addressing these limitations in future studies could help better understand IMAT accumulation and develop more effective interventions for improving sarcopenia in MHD patients.

In conclusion, we show that IMAT accumulation is significantly higher in patients with MHD than those without kidney disease. This accumulation is associated with metabolic abnormalities, body compositions, and physical impairments commonly observed in advanced CKD, suggesting that IMAT could play a crucial role in promoting muscle catabolism, which contributes to muscle weakness and functional decline. These findings underscore the importance of addressing IMAT accumulation as a potential intervention target to improve health outcomes and quality of life for individuals with advanced CKD.

## Data Availability

All data produced in the present study are available upon reasonable request to the authors.

