## Supplementary material for "INTERMUSCULAR ADIPOSE TISSUE AND MUSCLE FUNCTION IN PATIENTS ON MAINTENANCE HEMODIALYSIS": Figure S1

**MHD**

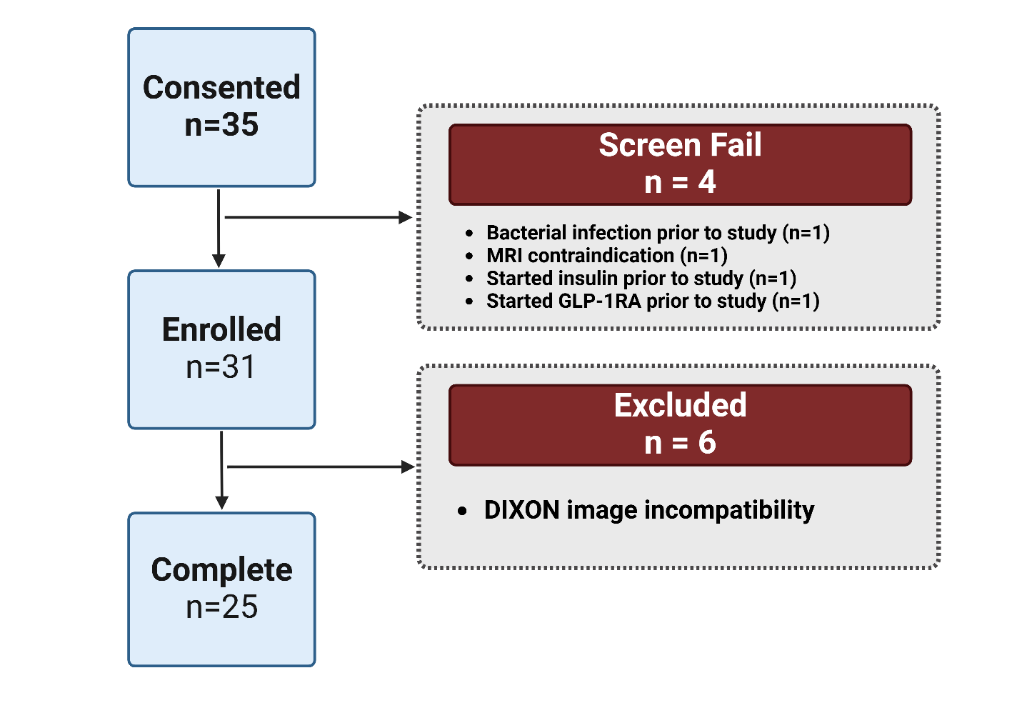

**Control**

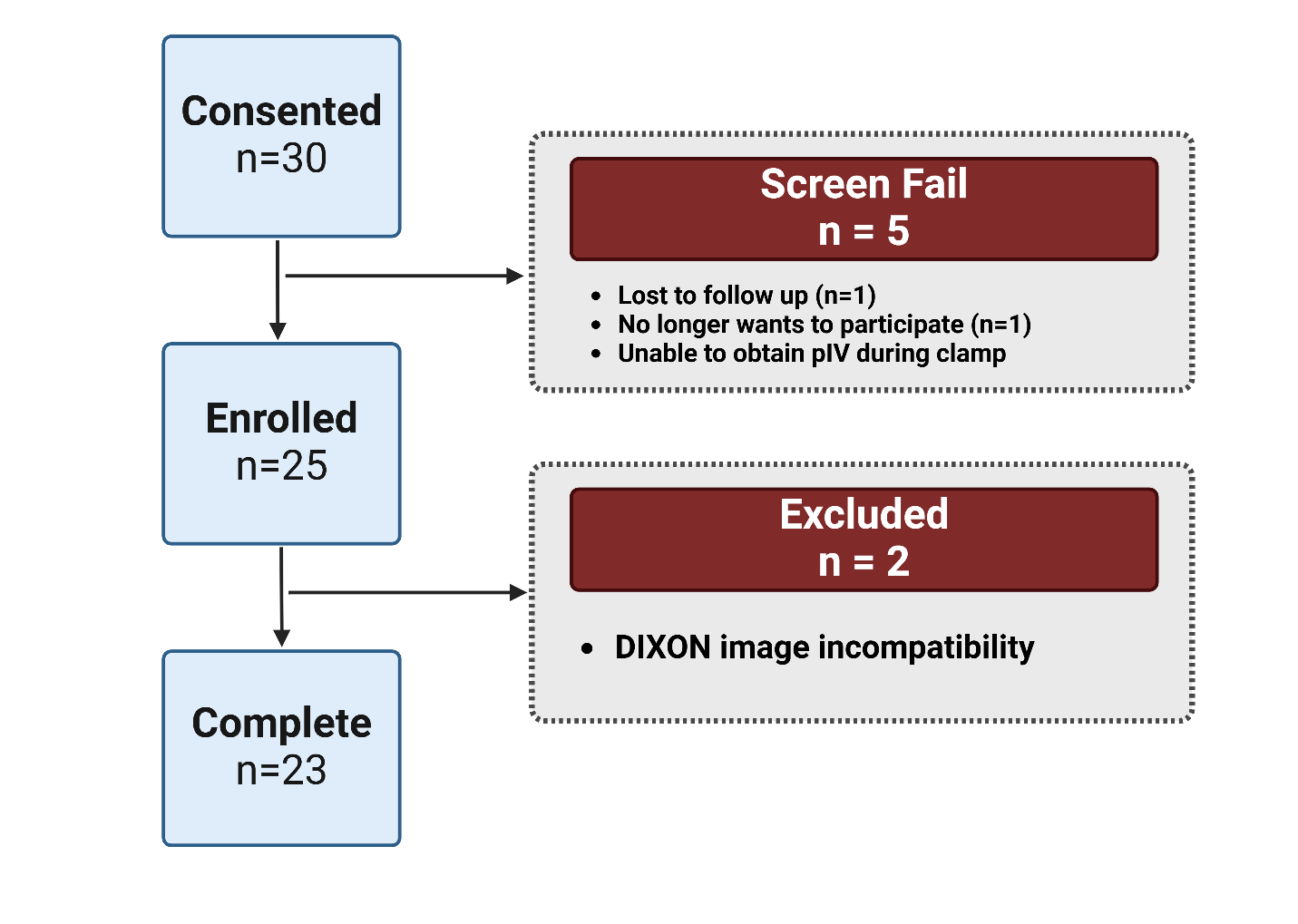

***Created with BioRender.com***

**Figure S1. Flow charts of MHD and Control groups respectively**
