## Supplementary material for "INTERMUSCULAR ADIPOSE TISSUE AND MUSCLE FUNCTION IN PATIENTS ON MAINTENANCE HEMODIALYSIS": Figure S2

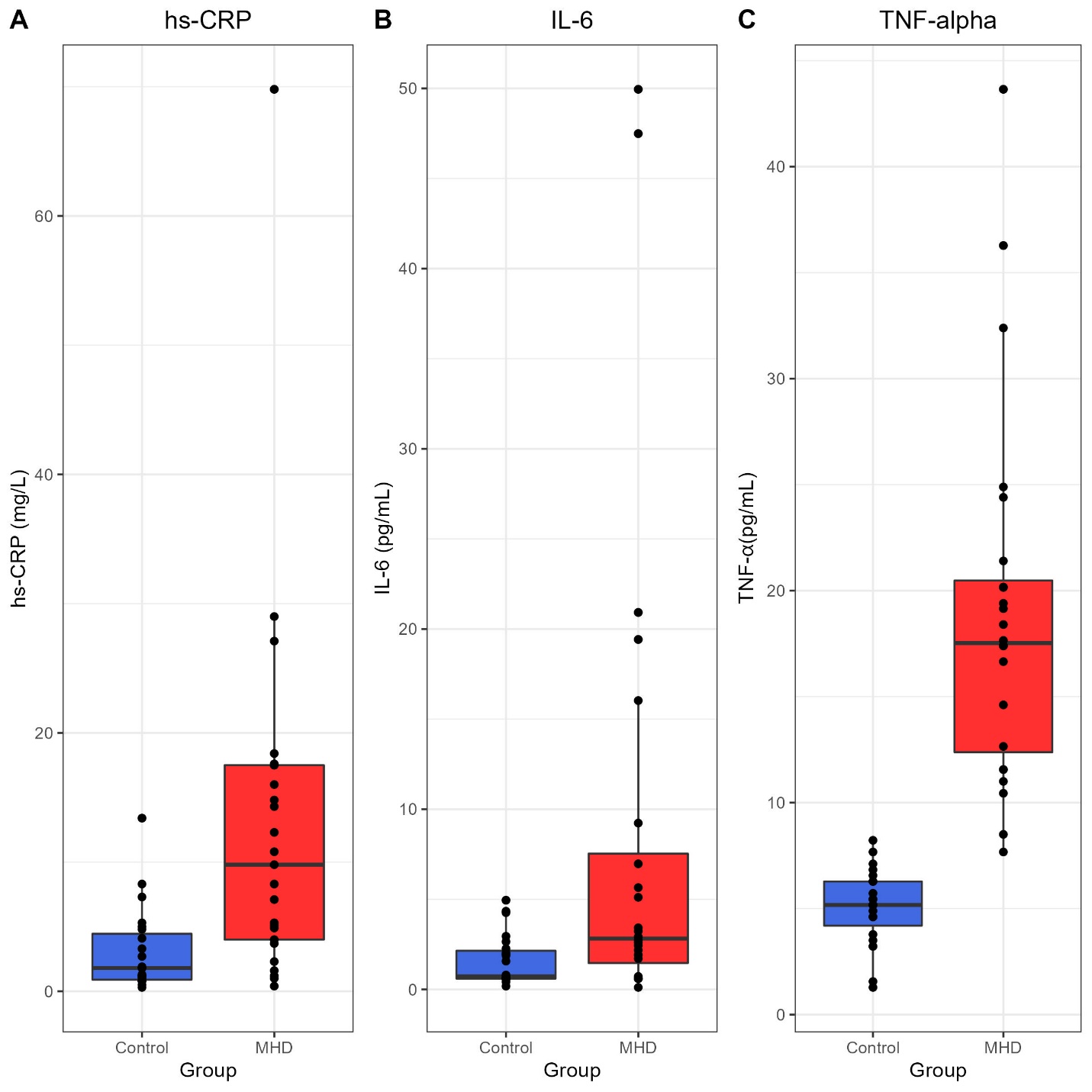

**Figure S2. Comparison of Inflammatory marker concentrations between groups.** **(A), (B), (C)** Box plot for TNF- α, IL-6, and hs-CRP concentrations comparison between control and MHD groups (p < 0.001, p = 0.004, and p < 0.001, respectively.)
