## Supplementary material for "INTERMUSCULAR ADIPOSE TISSUE AND MUSCLE FUNCTION IN PATIENTS ON MAINTENANCE HEMODIALYSIS": Table S1

**Table S1. Laboratory Values of Study Participants**

| Parameter | | Total (n = 49) | Control (n = 23) | MHD (n = 25) | P value |
| --- | --- | --- | --- | --- | --- |
|  | Hematocrit (%) | 41.0 (35.8 – 44.6) | 44.0 (41.5 – 46.0) | 36.0 (34.0 – 40.0) | P < 0.001* |
|  | Albumin (g/dL) | 4.0 (3.7 – 4.1) | 4.0 (3.8 – 4.2) | 3.9 (3.5 – 4.1) | p = 0.22 |
|  | Pre-albumin (mg/dL) | 26.0 (23.1 – 34) | 24.3 (22.1 – 26.0) | 33.5 (23.8 – 38.4) | p = 0.008* |
|  | Creatinine (mg/dL) | 6.11 (0.98 – 8.70) | 0.93 (0.81 – 1.03) | 8.66 (8.15 – 11.18) | p < 0.001* |
|  | Triglyceride (mg/dL) | 100 (66 – 128) | 100 (72- 109) | 101 (59 – 165) | p = 0.8 |
|  | Total Cholesterol (mg/dL) | 168 (128 – 191) | 180 (158 – 204) | 139 (118 – 182) | p = 0.009* |
|  | HDL Cholesterol (mg/dL) | 41 (32 – 46) | 43 (40 – 48) | 38 (30 – 42) | p = 0.007* |

Data are presented as median (IQR). *Indicates two-sided p ≤ 0.05 meets the significance criteria.
